# Knowledge, beliefs, practice challenges, training barriers, and needs of Kazakhstani nurses in the care of patients with autism: A descriptive cross-sectional study

**DOI:** 10.1101/2025.05.26.25328333

**Authors:** Akbota Tolegenova, Faye Foster, Akbota Kanderzhanova, Paolo Colet, Michelle Somerton, Valentina Stolyarova, J Gareth Noble

**Author notes:** **Corresponding author:** Akbota Tolegenova^1^, Address: NUSOM building, Kerey and Zhanibek Khans St, Astana 020000, Kazakhstan, **Email:**.

## Abstract

**Background:** Autism Spectrum Disorder (ASD) is a neurodevelopmental condition requiring early identification and intervention to improve outcomes. Nurses are key to ASD care, facilitating referrals and supporting interventions, but often face knowledge gaps and limited training. In Kazakhstan, rising diagnoses of ASD, and healthcare reforms highlight the need for nurse education, yet challenges remain underexplored. This study examines nurses’ knowledge, beliefs, challenges in practice, training barriers, and training needs in caring for patients with autism.

**Methods:** This cross-sectional descriptive survey assessed the knowledge, beliefs, challenges, training barriers, and training needs of nurses involved in the care of individuals with ASD in Kazakhstan. Data were collected using purposive sampling from nurses aged 18 years and older working in hospitals, schools, and autism centers through an online survey distributed via email, professional networks, and social media platforms. A bilingual online survey (Russian and Kazakh) included 52 questions on demographics, knowledge and beliefs, challenges, barriers, and training needs. Descriptive and bivariate analysis were conducted by using the R studio.

**Results:** The study included N=201 nurses, predominantly female (89.05%) with an average age of 37 years. Most worked in the capital city of Kazakhstan, Astana (66.67%) and government facilities (73.13%), with 66.17% having over five years of experience. Although 69.15% had encountered ASD cases, knowledge gaps were evident, particularly in recognizing traits like resistance to change (50.00%), symptom onset before age three (46.00%), and preoccupation with objects (49.75%). Nurses without formal ASD training had the lowest percentage agreement on true statements (45.30%) compared to those with formal education (61.40%) or professional development (60.10%). The support/school group showed the lowest understanding overall. Despite 91.04% expressing interest in training, barriers like time constraints (33.33%) and limited course availability (30.35%) highlight the need for structured educational training programs.

**Conclusion:** Nurses in Kazakhstan demonstrated interest in ASD training and face significant knowledge gaps, particularly among those without formal education or in support roles. While many have encountered ASD cases, misconceptions persist, underscoring the need for tailored training programs that address specific barriers such as time constraints and course availability. Therefore, we recommend a structured, accessible education initiatives that will be essential to enhance nurses’ understanding and capabilities in ASD care, ultimately improving patient outcomes.

**Clinical Relevance:** This study highlights significant knowledge gaps and training needs among Kazakhstani nurses in the care of patients with autism. Implementing structured, role-specific educational programs can improve early diagnosis, referral accuracy, and quality of care for individuals with ASD in clinical settings.

## Introduction

### Global Perspective on ASD

Autism spectrum disorder (ASD) is a complex neurodevelopmental condition characterized by challenges in social interaction, communication, and restricted behaviors (APA, 2013). Global estimates suggest that 1 in 100 children are diagnosed with ASD, with prevalence rates continuing to rise (Zeidan et al., 2022). This trend has sparked international calls for earlier identification and intervention— particularly in low- and middle-income countries (LMICs), where diagnostic infrastructure remains limited (Tasew et al., 2021; Danielson et al., 2022).

### ASD in Kazakhstan

ASD represents a relatively recent diagnostic concept in Kazakhstan, and the exact number of autistic children remains uncertain due to underdiagnosis and varying data sources. Nevertheless, official statistics reflect a steady increase in identified cases: 4,707 children were registered in 2018, 5,193 in 2019, 6,771 in 2020, 8,796 in 2021, and 12,087 by 2022 (Zhumagarin, 2022). This growth is attributed to greater public awareness and improvements in diagnostic practices.

### Healthcare Reforms and Challenges in Kazakhstan

In response, Kazakhstan’s healthcare system has begun implementing reforms to better support individuals with ASD, including the adoption of international diagnostic tools and the establishment of specialized autism centers (An et al., 2018). One significant development is the recent shift from ICD-10 to ICD-11, aligning Kazakhstan more closely with international standards (WHO, 2021). While countries like the United States adopted DSM-5 as early as 2013 (APA, 2013), and others, such as Italy, modified ICD-10 to incorporate DSM-5-like dimensions, Kazakhstan’s adoption of ICD-11 marks a critical step forward. Nonetheless, effective implementation will require focused training for healthcare professionals, particularly those at the primary care level.

### Role of Nurses in ASD Care

Nurses play an integral role within a multidisciplinary healthcare team, providing support throughout the diagnosis, assessment, and treatment processes (Flaubert et al., 2021). They are often the first point of contact for families, facilitating referrals for formal ASD evaluations by specialists such as developmental pediatricians or child psychologists and advocating for early intervention strategies tailored to each child’s unique need (Dunlap & Filipek, 2020). Additionally, nurses assist in implementing individualized interventions and therapies, ensuring comprehensive care that addresses both the behavioral and medical aspects of ASD (Flaubert et al., 2021. By leveraging evidence-based knowledge in screening and diagnostic methodologies, nurses are also pivotal in the early and accurate identification of ASD indicators. Furthermore, they provide essential hospital-based care and play a crucial role in supporting children and families through direct and frequent interactions within the healthcare system (Fraatz & Durand, 2021).

### Challenges Faced by Nurses

However, mounting evidence indicates that healthcare professionals, including nurses, often lack sufficient understanding of ASD, which can lead to delays in diagnosis and intervention (Gardner et al., 2016: Morris et al., 2019). Many nurses lack clinical experience and formal training on ASD, leading to significant knowledge gaps, particularly regarding its origins, comorbidities, and communication challenges (Ramu & Govindan, 2022; Corsano et al., 2019). Pediatric nurses especially report difficulties in communicating with children with ASD and often experience emotional strain such as sadness and inadequacy (Mahoney et al., 2021). These challenges highlight the urgent need for targeted education and training to improve nurses’ preparedness and confidence in supporting individuals with ASD.

Alongside the clinical and educational challenges, this study also recognizes the emotional and ethical realities of nursing work. Nurses do more than provide care and they often carry the emotional weight of their patients and navigate ethical tensions, especially when systems fail to support them. As Tronto (1993) explains, good care involves being attentive, responsible, competent, and responsive, but these qualities are difficult to maintain without proper training or resources. Hochschild (1983) also reminds us that nurses engage in emotional labor, constantly managing their own feelings while offering support to patients and families. These perspectives help us better understand the everyday struggles Kazakhstani nurses face when caring for children with autism—struggles that go far beyond clinical checklists or protocols.

### Systemic Issues and Training Needs

Beyond knowledge gaps, systemic issues further complicate ASD care in Kazakhstan. These include the absence of specialized nursing roles for autism, a lack of standardized training programs, limited interdisciplinary collaboration, and insufficient educational resources. These barriers are exacerbated by widespread stigma and a shortage of targeted training opportunities (An et al., 2018; Somerton et al., 2022). Despite the urgency of the issue, research on healthcare providers’ knowledge, attitudes, and practices remains scarce—especially among nurses. Consequently, there is limited understanding of the specific challenges they face or the training they require to effectively support individuals with ASD and their families.

### Research Gaps and Study Objectives

Recent studies have begun exploring the challenges faced by Kazakhstani healthcare providers in ASD diagnosis and care. For example, Somerton et al. (2022) identified persistent gaps in understanding the barriers that health professionals encounter, calling for more research into training and systemic support needs. The only known study focused specifically on Kazakhstani nurses’ awareness of ASD conducted by Kosherbayeva et al. (2024) revealed concerning gaps: while 70.9% of nurses were aware of the ICD-11 update, only 41.7% knew all ASD diagnostic methods, and just 27.8% were familiar with the modified autism screening test. Alarmingly, 66.7% misclassified ASD as a mental disorder, and only 9.1% expressed interest in further education. Pediatric nurses demonstrated slightly better knowledge compared to general nurses. Moreover, nurses, who form the backbone of Kazakhstan’s healthcare workforce, are rarely included in targeted ASD training or policy development, despite frequent clinical exposure to autistic patients (Tolegenova et al., 2024). Parallel findings from caregiver research in Kazakhstan show that mothers of autistic children also shoulder disproportionate emotional and logistical burdens within similarly fragmented systems (Foster et al., 2025).

### Study Aim

Therefore, this study aims to address these gaps by conducting a comprehensive needs assessment to explore the nurses’ knowledge, beliefs, challenges in practice, training barriers, and training needs in caring for patients with autism. Furthermore, the findings of this research will inform the development of a professional development program to address the needs identified by this research.

### Study Design

This study employed a cross-sectional descriptive survey design to capture a broad snapshot of nurses’ knowledge, beliefs, experiences, and training needs related to ASD in Kazakhstan. This design was selected for its suitability in efficiently assessing a large and diverse sample across multiple healthcare settings.

## Materials & Methods

The target population consisted of nurses actively involved in the initial screening, assessment, referral, treatment, or management of children with ASD in urban areas of Kazakhstan. Inclusion criteria required participants to be nurses aged 18 years or older with professional experience in healthcare settings such as hospitals, schools, or autism centers. Nurses who were not engaged in ASD-related care or diagnosis, or who were unable to provide informed consent, were excluded from participation. A non-probability sampling approach was used, specifically a combination of convenience and voluntary response sampling. The survey was disseminated by clinics and hospitals through professional networks and social media platforms. As the research team did not directly control distribution or have access to information about the total number of nurses invited, it was not possible to calculate a formal response rate.

### Instrument Construction

Data were collected between June-August of 2024 using a self-administered online survey developed via SurveyMonkey, available in both Russian and Kazakh. The instrument consisted of 52 items divided into four key sections:

#### Sociodemographic information

Included questions on age, gender, clinical experience, workplace location, and professional roles.

#### Knowledge and beliefs

This section was adapted from the work of Imran et al. (2011) and included structured items where nurses categorized various behaviors as “Necessary,” “Helpful but Not Necessary,” or “Not Helpful” for autism diagnosis. For example, lack of social responsiveness and language delay were expected to be rated as “Necessary,” while restricted interests were rated as “Helpful but Not Necessary.” Incorrect classifications—such as rating lack of eye contact as “Not Helpful”—were used to identify knowledge gaps. Additionally, this section featured belief-based 21 statements regarding the nature, causes, prognosis, and treatment of ASD, rated on a 3-point Likert scale: Agree, Not Sure, or Disagree. The items were designed to capture both accurate beliefs (e.g., “Autism is a developmental disorder”) and common misconceptions (e.g., “Autistic children usually grow up to be schizophrenic adults”), allowing the identification of both knowledge strengths and gaps.

#### Barriers and challenges

Consisted of criterion-based questions about the challenges nurses face in identifying and managing ASD, including limited resources and systemic constraints.

#### Training needs

Addressed previous training, interest in further education, preferred formats (e.g., webinars, online self-paced), and priority topics like diagnosis or patient management.

The survey also incorporated open-ended questions (e.g., *“Are you aware of any specialized center for autism in Kazakhstan?”, “Where would you refer a child if you diagnose or suspect autism?”*, and *“What other comments would you like to add about your experiences with autism?”*) to gain qualitative insights into participant perspectives. Content validity was supported through expert review by three professionals in nursing and public health.

The survey was first developed in English and then translated into Russian and Kazakh by bilingual experts. A back-translation process was conducted to confirm accuracy and consistency with the original version. Discrepancies were reviewed and resolved collaboratively by the translation team. The survey underwent pilot testing with a small sample of five nurses (n = 5) to assess clarity and flow.

Feedback from the testing indicated that the instrument was clear and relevant, and no substantial changes were made. While no formal reliability testing was conducted, expert and participant feedback supported the tool’s face and content validity. Participants were recruited through purposive sampling using targeted emails, professional networks, and social media platforms (WhatsApp, Telegram, and Instagram).

### Data Analysis

#### Quantitative data were analyzed using statistical software R studio version 3.6.3 (2020-02-29)

The study employed descriptive statistics, comparative analysis, and thematic analysis. Descriptive statistics summarized the socio-demographics and clinical experience of participants, while comparative analysis examined differences in knowledge and beliefs of participants based on training and area of specialization. Data were also grouped by training formats and specializations, to understand a comprehensive view of nurses’ knowledge and beliefs and areas requiring enhanced ASD education.

For further analysis the instruments recorded the previous training of participants by type and these were grouped as shown below:

1. Formal education (includes structured learning that is part of a formal education system, typically completed during a degree or certification): During college, Lectures.
2. Professional development (includes structured training for healthcare professionals, such as courses, conferences, and continuing medical education): Professional continuing medical education training, Conferences, Specific autism course/training.
3. Self-led learning (includes learning driven by the individual’s own initiative, not formally organized by an institution): Self-education, Webinars.
4. No formal training: Not Attended.

The average was calculated by adding all the values for Agree and Not sure/Disagree for each category separately and divided by number of statements.

Next the Specializations were also grouped as following according to their functions:

1. Management: Chief Nurse, Senior Nurse
2. Clinical: Advanced Practice Nurse, District Nurse, General Practice Nurse, Feldsher
3. Support/School: Specialized Nurse, School Nurse in Educational Institutions, Junior Nurse.

For qualitative data, responses to the open-ended survey questions were analyzed using thematic analysis to identify recurring patterns and key areas of concern. An inductive approach was used, allowing themes to emerge directly from the data without imposing pre-existing frameworks. First, all responses were read multiple times for familiarization. Initial codes were then generated manually by identifying phrases and statements that reflected common ideas or concerns. These codes were grouped into broader thematic categories based on conceptual similarity. The process was iterative, with themes refined through constant comparison and cross-checking. Four dominant themes were identified: challenges in recognizing and managing autism, need for training and education, lack of systemic resources and support, and low public and institutional awareness. This qualitative analysis enriched the quantitative findings by capturing nurses’ personal perspectives and highlighting contextual challenges not easily measurable through structured questions.

### Ethical Considerations

The study was conducted in accordance with Nazarbayev University Institutional Research Ethics Committee code of ethics to ensure participant rights and confidentiality. Study invitations provided detailed information and emphasized voluntary participation. Informed consent was obtained electronically prior to survey participation. Data were anonymized, securely stored, and accessible only to authorized research personnel. Participants were informed of their right to withdraw them or their data from the study at any point without any repercussions.

## Results

### Socio-Demographics of Participants

This study involved N=201 nurses, mainly female (89.05%), averaging 37 ± 12.25 years old. Most are based in the capital of Kazakhstan, Astana (66.67%) and work primarily in government facilities (73.13%) (Table 1). The largest professional groups are Advanced Practice Nurses (21.89%) and District/General Practice Nurses (20.40%). A majority have over five years of experience (66.17%), and 69.15% have encountered ASD cases in their clinical work.

**Table 1.**
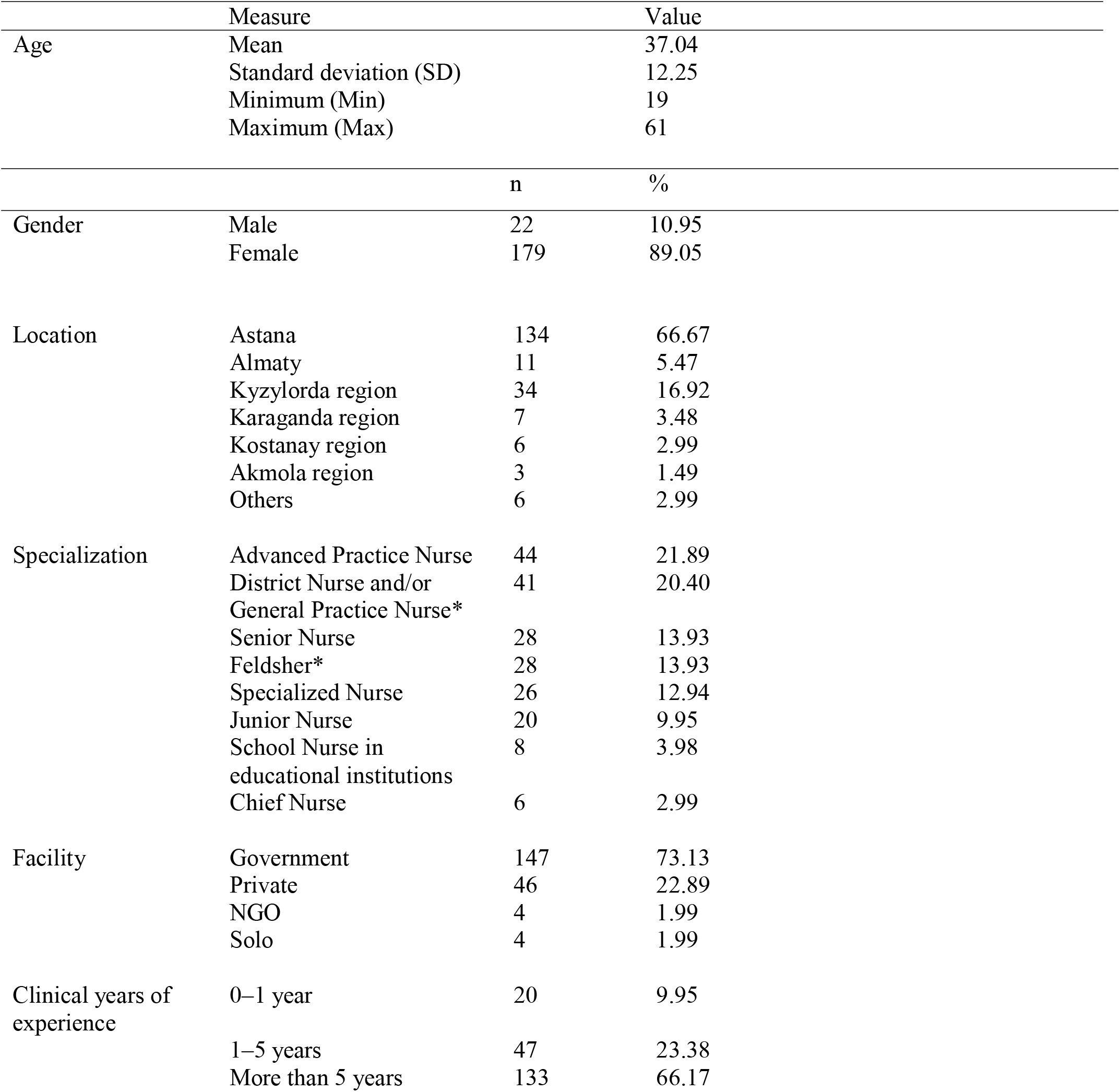
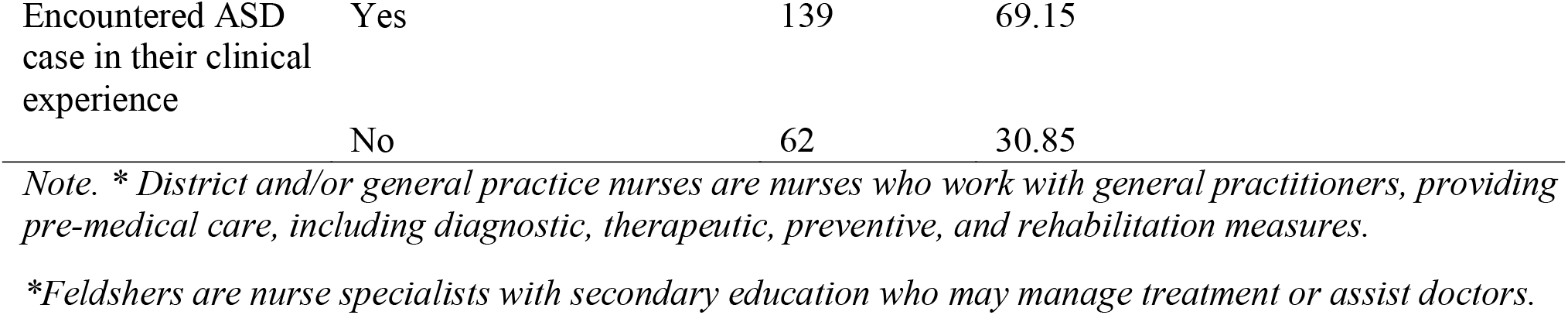
Socio-Demographics of Participants (N = 201)

### Knowledge and Beliefs Overall

Nurses’ knowledge about the general characteristics required for a diagnosis of individuals with ASD was assessed in the survey based on common characteristics and were criterion based. For example, ‘essential’, ‘necessary’, etc. These data show language delays (69.65%), peculiar speech characteristics (66.67%), and social interaction difficulties (63.18%) were the most often rated as “necessary”. Some traits are seen as “helpful but not necessary,” such as resistance to change in routine (43.28%) and onset of symptoms before 36 months (45.27%). Finally, a few characteristics were rated as “not helpful.” The highest percentages in this group were for unusual mannerisms (11.94%) and preoccupation with objects (11.94%) (see Supplement 1).

Nurses’ beliefs about ASD were assessed through 22 statements rated on a 3-point Likert scale (Agree, Not Sure, Disagree), capturing both accurate views and common misconceptions to identify belief patterns and gaps. The results revealed an agreement for statements like “Autism can occur in mild as well as extreme forms” (63.18%), “Autism is a communication disorder” (66.17%), and “Autism is a developmental disorder” (60.70%). Uncertainty was most evident for statements such as “Autism exists only in childhood” (34.83% unsure), “Autistic children do not show social attachments” (51.74% unsure), and “Autism is a lifelong condition” (49.25% unsure). Disagreement percentage was highest for “Autistic children usually grow up to be schizophrenic adults” (28.36% disagree) and “Autism is an emotional disorder” (8.46% disagree).

Speech therapy (20.98%) and special education (20.33%) were the most commonly selected interventions, indicating strong recognition of these supporting interventions. There is a general understanding among nurses of therapeutic and educational interventions for individuals with ASD, however it also shows some inclination toward pharmacological treatments. Six participants replied that they do not know by choosing the “Other” option (see Supplement 1).

To see training background’s influence on knowledge accuracy average was calculated by adding all the values for Agree and Not sure/Disagree for each category of previous training separately and divided by number of true statements and misleading statements. From the results it can be seen that training background influenced knowledge accuracy. Nurses with formal education had the highest agreement with true statements (61.4%), followed by those in professional development (60.1%) and self-education (56.0%). Those without formal training had the lowest average (45.3%). Misconception resistance was strongest among formal education (75.1% not sure/disagree), though professional development participants showed more agreement with misleading statements (35.3%).

The categorized analysis of knowledge by specialization type highlighted differences in recognizing ASD characteristics across clinical, management, and support/school groups. By specialization, clinical nurses demonstrated the strongest recognition of core traits like language delays (73.0%) and social interaction difficulties (62.0%). Management nurses followed closely, while support/school nurses had the lowest “necessary” ratings, indicating a knowledge gap in that group (see Supplement 1).

### Awareness of ASD Resources and Referral Practices

Findings revealed inconsistent knowledge about autism resources and referral pathways. The most recognized center was Asyl Miras (*n*=47), followed by Orda (*n*=22) and the National Center for Pediatric Rehabilitation (*n*=16). The University Medical Center (UMC) was mentioned only once. Notably, 56 participants were unaware of any specific autism centers, highlighting a significant knowledge gap. Other facilities, such as the Rehabilitation Center (*n*=6), Autism Center in Almaty (*n*=3), and Golden Kids (*n*=2), were mentioned occasionally.

When asked where they would refer a child suspected of autism, most participants cited psychiatrists (*n*=37), followed by psychologists (*n*=17), neurologists (*n*=16), general practitioners or therapists (*n*=13), and neuropathologists (*n*=12). Less common referrals included pediatricians, speech therapists (*n*=3), neuropsychologists, psychotherapists (*n*=2 each), and defectologists or surdologists (*n*=1 each). Notably, 22 participants were unsure where to refer a child.

### Training Experiences and Barriers

Among the N=201 participants, 50.75% reported difficulties in identifying or recognizing ASD, while 31.34% stated they did not face such barriers (Table 2). Regarding challenges in nursing practice (N=379), the most cited issues were a lack of knowledge on ASD signs and symptoms (19.00%), followed by a combination of multiple barriers (15.04%), and limited access to ASD resources (13.19%). Additional concerns included the absence of guidelines for nursing care (11.61%) and insufficient in-service training (8.44%).

**Table 2.** Nurses’ previously attended trainings and barriers in practice.

|  |  | n | % |
| --- | --- | --- | --- |
| Face difficulties or barriers in identifying and/or recognizing ASD (N = 201) | Yes | 102 | 50.75 |
|  | No | 63 | 31.34 |
|  | Not applicable | 36 | 17.91 |
| Challenges and Barriers in nursing practice (N total= 379) | Lack of knowledge on ASD/ASD criteria, signs and symptoms | 72 | 19.00 |
|  | All of the above | 57 | 15.04 |
|  | Lack of knowledge about ASD resources (books, websites) | 50 | 13.19 |
|  | Lack of guidelines for the nursing care and management of a pediatric patient with ASD | 44 | 11.61 |
|  | Lack of in-service or continuing education (training) on ASD | 32 | 8.44 |
|  | Lack of understanding of language development and communication in ASD | 30 | 7.92 |
|  | Parental knowledge of the patient's individual needs and routines | 30 | 7.92 |
|  | None of the above | 30 | 7.92 |
|  | Lack of Kazakh/Russian adaptations of tools | 25 | 6.60 |
|  | Not enough support from administration | 6 | 1.58 |
|  | Other | 3 | 0.79 |
| Previous training type<br>(N total= 201) | Not attended | 94 | 46.77 |
|  | Self-education | 27 | 13.43 |
|  | Specific autism course/training | 23 | 11.44 |
|  | Professional continuing medical education training | 18 | 8.96 |
|  | Conferences | 18 | 8.96 |
|  | Webinars | 8 | 3.98 |
|  | During college | 7 | 3.48 |
|  | Lectures | 6 | 2.99 |
| Previous training topics<br>(N = 201) | Not attended | 94 | 46.77 |
|  | Screening/diagnostics | 75 | 37.31 |
|  | Treatment/therapy | 32 | 15.92 |

Regarding the previous training, 46.77% of nurses indicated that they had not attended any ASD-related training. Of those who had training, self-education (13.43%) and specific autism courses (11.44%) were the most common forms, while professional education programs and conferences were less frequent. Training topics focused primarily on screening and diagnostics (37.31%), with less emphasis on treatment and therapy (15.92%).

The results for training interest show strong interest among nurses (91.04%) in attending training on ASD, with only a small percentage (8.96%) not interested (Table 3). Key barriers to participation include lack of time (33.33%), limited availability of ASD courses (30.35%), and insufficient information on available courses (19.40%).The areas nurses are most interested in learning about are Patient care (24.88%), Diagnostics (23.38%), Patient communication (16.92%), and Screening (12.44%). Regarding training formats, online self-directed learning (videos and reading materials) is the most favored (35.32%), followed by online interactive webinars (26.37%). In-person seminars in Astana or with traveling lecturers are less popular (9.95% and 3.98%, respectively), while 23.38% of nurses expressed no specific preference.

**Table 3.**
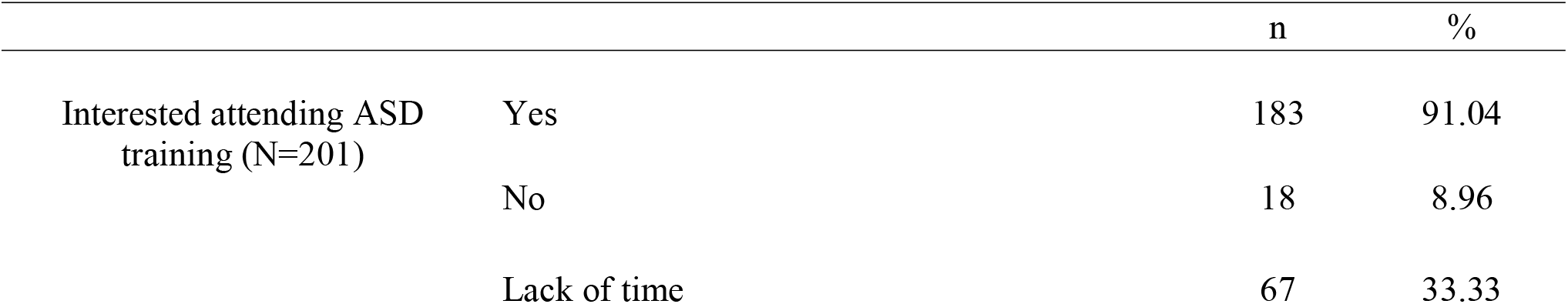
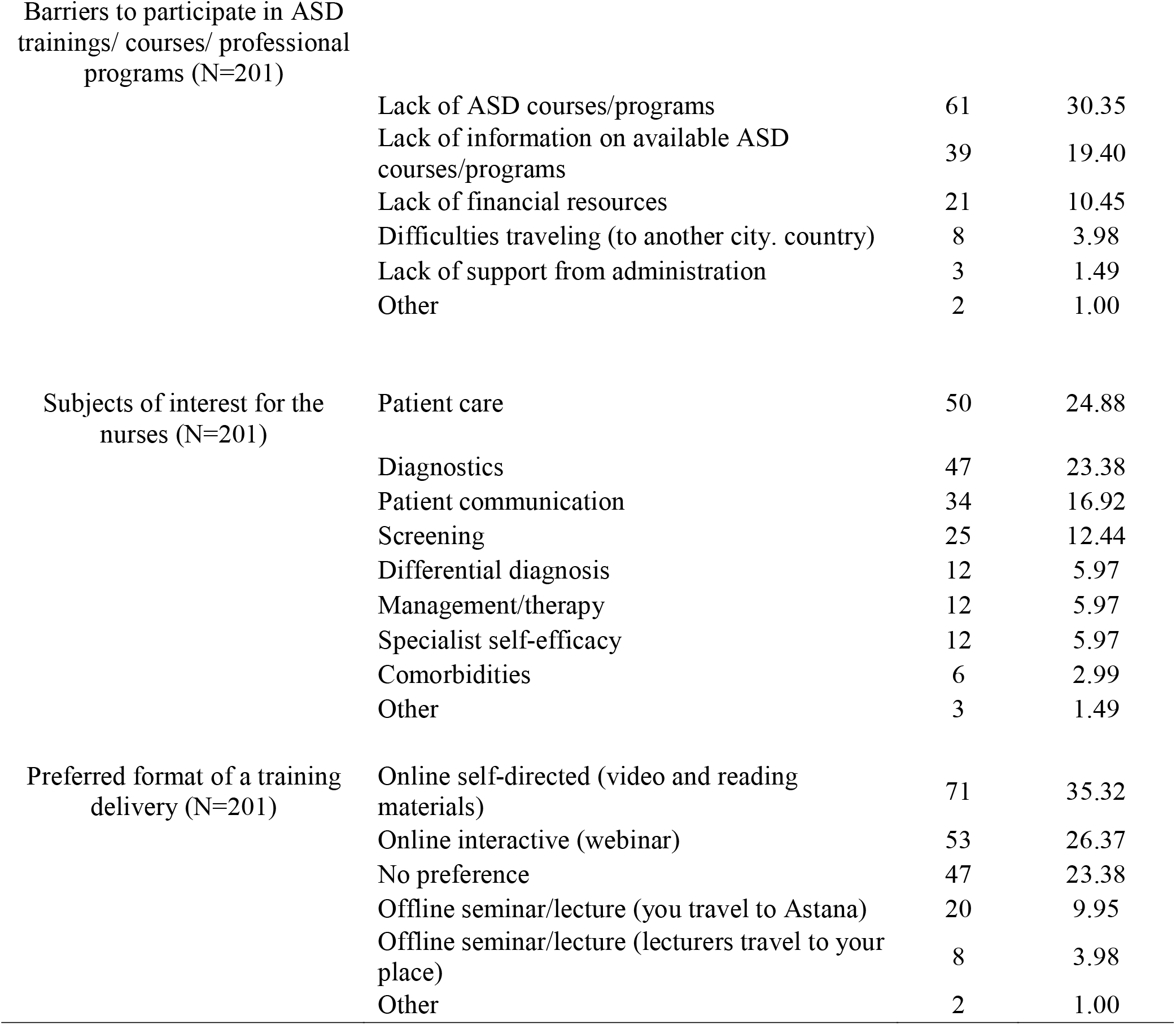
Training needs of nurses in ASD management.

|  |  | n | % |
| --- | --- | --- | --- |
| Interested attending ASD training (N=201) | Yes | 183 | 91.04 |
|  | No | 18 | 8.96 |
|  | Lack of time | 67 | 33.33 |

Barriers to participate in ASD trainings/ courses/ professional programs (N=201)
|  |  |  |
| --- | --- | --- |
| Lack of ASD courses/programs | 61 | 30.35 |
| Lack of information on available ASD courses/programs | 39 | 19.40 |
| Lack of financial resources | 21 | 10.45 |
| Difficulties traveling (to another city. country) | 8 | 3.98 |
| Lack of support from administration | 3 | 1.49 |
| Other | 2 | 1.00 |

Subjects of interest for the nurses (N=201)
|  |  |  |
| --- | --- | --- |
| Patient care | 50 | 24.88 |
| Diagnostics | 47 | 23.38 |
| Patient communication | 34 | 16.92 |
| Screening | 25 | 12.44 |
| Differential diagnosis | 12 | 5.97 |
| Management/therapy | 12 | 5.97 |
| Specialist self-efficacy | 12 | 5.97 |
| Comorbidities | 6 | 2.99 |
| Other | 3 | 1.49 |

Preferred format of a training delivery (N=201)
|  |  |  |
| --- | --- | --- |
| Online self-directed (video and reading materials) | 71 | 35.32 |
| Online interactive (webinar) | 53 | 26.37 |
| No preference | 47 | 23.38 |
| Offline seminar/lecture (you travel to Astana) | 20 | 9.95 |
| Offline seminar/lecture (lecturers travel to your place) | 8 | 3.98 |
| Other | 2 | 1.00 |

### Qualitative Insights from Open-Ended Responses

Thematic analysis of open-ended responses (n = 201) revealed four interconnected themes: (1) Clinical Confusion and Recognition Challenges, (2) High Demand for Training, (3) High Demand for Training, and (4) Lack of Resources and Support Infrastructure.

1. Clinical Confusion and Recognition Challenges: Many nurses expressed difficulty understanding and recognizing ASD symptoms, with comments such as “It is very difficult to understand such children” and “Could not recognize the patient.” These remarks align with the survey’s quantitative findings, where over 50% reported uncertainty about key ASD traits (e.g., social attachments, prognosis). This suggests a gap between textbook definitions and real-world recognition, highlighting a need for experiential or simulation-based training.
2. High Demand for Training: There was a clear and recurring emphasis on education: “It is necessary to get a comprehensive education”, “Continuous training”, and “Parents should be educated about ASD.” This eagerness complements the 91% of participants who expressed interest in ASD training in the quantitative section. Importantly, some comments advocated for training for both professionals and families, underscoring a system-wide educational gap.
3. Lack of Resources and Support Infrastructure: Respondents called for more autism centers, trained specialists, and logistical support: “We need more centers”, “Open more schools”, and “Create a center to support mothers who have autistic children.” This reflects broader systemic issues in Kazakhstan’s ASD care landscape and supports calls from An et al. (2018) and Somerton et al. (2022) for expanded autism-specific services and professional roles.
4. Societal and Institutional Awareness Gaps: A final theme was concern about low public and institutional awareness: “The population is not aware of autism”, “There are no specialized, interested specialists”, and “This problem is not being solved at the state level.” These statements emphasize both social stigma and systemic inertia. Nurses also noted the financial burden families face, revealing how socioeconomic pressures intersect with clinical care.

## Discussion

This study represents the first large-scale examination of Kazakhstani nurses’ knowledge, beliefs, needs and barriers in caring for patients with ASD. Although the majority of nurses reported clinical contact with patients on the spectrum, the findings reveal considerable knowledge gaps, widespread misconceptions, and significant systemic and educational barriers. These results echo findings from other LMICs and reinforce the urgent need for context-specific training and support mechanisms.

### Foundational Knowledge and Misconceptions

Nurses demonstrated a basic understanding of core ASD traits, with high recognition of language delays, peculiar speech, and social difficulties. However, understanding of understated indicators, such as preoccupation with objects and unusual mannerisms was limited, with 11.94% rating them as “not helpful”. While a majority acknowledged autism as a developmental and communication disorder, misconceptions about its prognosis persisted. Nearly half (49.25%) were unsure if autism is lifelong, and 43.28% were uncertain whether it could be outgrown. These findings align with Somerton et al. (2022), where 29% (13 out of 45) of health specialists from Kazakhstan agreed that autism can be outgrown, and another 44% were unsure. Misconceptions about the permanence of ASD may contribute to delays in referral and support, particularly for children and adults. Training programs must explicitly counter these myths using real-world case examples and lifespan-focused content.

In a recent study, 54% of nurses misclassified autism as an emotional disorder, and 18% believed that autistic children might develop schizophrenia in adulthood. These misconceptions are consistent with findings from other LMICs, where autism is frequently misinterpreted as a psychiatric or emotional disorder. For instance, a 2019 study in Saudi Arabia revealed that 72.1% of healthcare professionals considered medication as the primary treatment for autism, and 38.1% were skeptical about the benefits of early interventions, indicating a limited understanding of ASD management (Hayat et al., 2019).

Similarly, a 2020 study in Sri Lanka found that 43.6% of public health midwives believed that poor parental attention caused autism, and 42.2% attributed it to parental conflicts during pregnancy and early childhood (Rohanachandra et al., 2020). Such misunderstandings not only misinform clinical diagnoses but might also lead to stigmatization, delayed intervention, and inappropriate treatment choices. There is a clear need for training programs to differentiate ASD from emotional and psychotic disorders by incorporating case studies and diagnostic criteria into curricula, as emphasized in recent regional training reviews that call for integrated, interdisciplinary curricula to address conceptual confusion and build diagnostic confidence across health professions (Foster et al., 2025)

Knowledge levels differed across nursing roles. Clinical nurses showed the strongest understanding of diagnostic traits, particularly language and social delays, while school nurses exhibited lower recognition. This variation highlights the importance of role-specific training tailored to the responsibilities and patient interactions of different nurse categories (Danielson et al., 2022).

Additionally, experience alone was not a guarantee of knowledge: while 66.17% had over five years of clinical work, knowledge accuracy remained tied more closely to formal training than to the years in practice.

### Training Gaps and Educational Needs

Despite strong interest in further training (91.04%), nurses reported key barriers such as time constraints (33.33%), limited course availability (30.35%), and insufficient information about existing opportunities (19.40%). These findings are consistent with a study by Alruwaili et al. (2023), which found that pediatric nurses in Saudi Arabia stated limited access to training resources and lack of time as significant barriers to ASD education. Additionally, there was a high percentage of preference among nurses for flexible, accessible learning formats, favoring online self-directed modules (35.32%) and webinars (26.37%), a trend also noted in other researches (Shawahna, 2021).

The challenges described by participants, particularly emotional strain, diagnostic uncertainty, and lack of systemic support, can also be viewed through a broader ethical and emotional lens. According to Tronto’s ethics of care (1993), effective care requires attentiveness to needs, competence in delivery, and responsiveness from institutions, all of which appear insufficient in the current system. Many nurses expressed frustration over the lack of tools and institutional guidance, which may hinder their ability to fully engage in this moral process. Furthermore, the emotional exhaustion described by some participants reflects what Hochschild (1983) termed emotional labor, where professionals must regulate their feelings while maintaining a caring façade. In the context of ASD, where diagnosis is complex and public understanding is limited, this emotional burden is intensified. Recognizing these dimensions supports the argument that nurses not only need technical knowledge, but also institutional structures that value and support the emotional and ethical demands of their roles. These interconnected dynamics are visually represented in Figure 1, which illustrates how knowledge gaps, misclassifications, and practice challenges intersect to shape training needs and care outcomes. The model also integrates Tronto’s and Hochschild’s frameworks to highlight how ethical care and emotional labor are embedded in nurses’ daily experiences. By using this theoretical lens, the study underscores the need for ASD training programs that are not only clinically informative but also responsive to the real-world emotional and moral pressures nurses face in practice.

**Figure 1.**
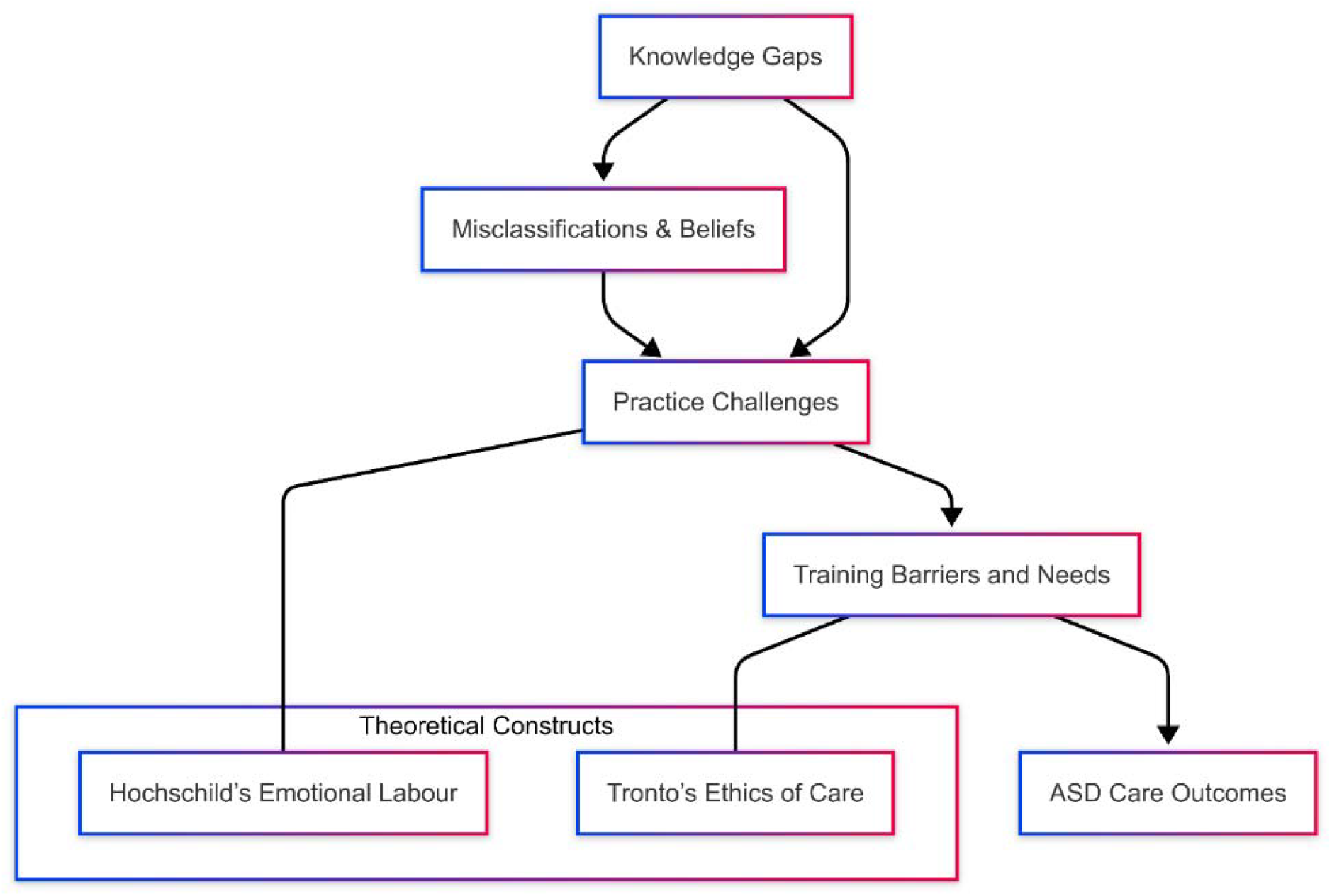
Conceptual Framework: Interrelationship Between Knowledge Gaps, Practice Challenges, and ASD Care Outcomes Through a Theoretical Lens

This model illustrates how nurses’ knowledge gaps and misclassifications contribute to practice challenges in ASD care (Figure 1). These challenges are further shaped by training barriers and needs, which ultimately influence care outcomes. Hochschild’s theory of emotional labor and Tronto’s ethics of care offer insight into the systemic and emotional dimensions of these experiences.

### Referral Practices and Systemic Challenges

Referral practices varied widely, with 22 respondents unsure where to refer suspected ASD cases. This suggests inconsistencies in clinical guidance, which may hinder early intervention efforts. Given that early diagnosis is critical for effective intervention (Dunlap & Filipek, 2020), addressing these gaps is imperative. Additionally, many nurses were unaware of autism centers in Kazakhstan, and unclear referral processes persisted, further complicating care access. Strengthening knowledge of available resources and establishing clear referral protocols would enhance support for autistic individuals and their families nationwide (Tolegenova et al., 2024). The findings also underscore the critical importance of ongoing education and professional development for nurses. To address evolving care needs, regular in-service training and continuing education programs focused on ASD are essential. These initiatives enable nurses to stay up-to-date with emerging research and best practices, thereby enhancing the quality of care provided (Shawahna, 2021; Tolegenova et al., 2024). However, this goal is difficult to achieve without strong institutional support. Notably, a low percentage of nurses reported receiving adequate backing from their administration, revealing a significant gap. By strengthening this support, healthcare institutions can help ensure more effective and consistent ASD care, ultimately leading to better outcomes for patients and their families (Fraatz & Durand, 2021). In addition, the lack of diagnostic and training tools in Kazakh and Russian was found to create a major barrier. As highlighted by Zakirova-Engstrand & Yakubova (2023), culturally and linguistically adapted materials are essential for effective learning and patient communication.

### Qualitative Insights and Broader Systemic Issues

Thematic analysis of open-ended responses revealed four interconnected concerns: clinical uncertainty, a strong desire for education, lack of systemic support, and limited public awareness. Nurses expressed difficulty in recognizing and communicating with ASD patients, a strong need for continuing education, and frustration with the limited number of diagnostic and treatment centers. Some emphasized the absence of state-level solutions, the rising number of ASD cases, and the high cost of private services. These perspectives not only validate the quantitative findings but also underscore the emotional burden and systemic constraints that limit nurses’ capacity to provide effective care. There is a clear need to increase institutional support, expand service access, and improve nursing education on ASD. These insights are consistent with findings from Somerton et al. (2022) and Kosherbayeva et al. (2024), which similarly identified gaps in institutional support, a lack of interdisciplinary collaboration, and insufficient ASD training for healthcare professionals across Kazakhstan. Together, these findings highlight the urgent need to expand service access, improve nursing curricula, develop interdisciplinary training, and ensure institutional and governmental backing to improve ASD care at all levels.

### Limitations and Future Research

Several limitations must be considered. The sample was limited to nurses with internet access and primarily urban, government-employed respondents, reducing generalizability to rural and private healthcare settings. Self-reported data may also be affected by social desirability bias. Future studies should explore how nurses apply ASD-related knowledge in clinical settings and evaluate the effectiveness of training interventions over time. Longitudinal research tracking changes in confidence, diagnostic accuracy, and caregiver satisfaction would provide valuable insights into the impact of professional development programs. Additionally, observational or ethnographic studies could examine how nurses navigate systemic barriers and collaborate with other professionals in real-world settings.

Comparative research across regions or between urban and rural facilities could further illuminate disparities in ASD care access and support.

### Recommendations for Practice and Policy

To address the identified gaps, several key actions are recommended. Develop culturally and linguistically appropriate ASD training programs in Kazakh and Russian to ensure accessibility. Integrate ASD-focused modules into nursing curricula and continuing education to build foundational knowledge. Training should prioritize myth-busting and enhancing diagnostic confidence. Strengthen interdisciplinary collaboration and improve awareness of available autism resources. Finally, ensure strong administrative support to enable consistent access to training, clinical guidelines, and necessary infrastructure.

## Conclusion

This study reveals important knowledge and system-level gaps in ASD care among Kazakhstani nurses. Despite strong interest in training and foundational awareness of ASD, misconceptions, inconsistent referral knowledge, and limited institutional support hinder effective practice. Addressing these challenges through structured, role-specific, and linguistically accessible training programs, supported by institutional commitment, will be vital for improving early diagnosis and care outcomes for individuals with ASD across Kazakhstan.

### Clinical Resources

- **Autism Speaks - Toolkits for Health Professionals** https://www.autismspeaks.org/tool-kits Offers free downloadable toolkits on screening, diagnosing, and managing ASD, tailored for healthcare providers.
- **Centers for Disease Control and Prevention (CDC) – Autism Spectrum Disorder (ASD)** https://www.cdc.gov/ncbddd/autism Provides current information on ASD signs and symptoms, diagnosis, screening guidelines, and training tools.
- **World Health Organization (WHO) – Autism Spectrum Disorders** https://www.who.int/news-room/fact-sheets/detail/autism-spectrum-disorders Includes global data on prevalence, challenges in care, and recommendations for health systems in LMICs.

## Data Availability

All data produced in the present study are available upon reasonable request to the authors

## Acknowledgement

This work was supported by Nazarbayev University (Faculty-Development Competitive Research Grants Program, 2023–2025, Project #20122022FD4132).

## Notes

**Declaration of conflicting interests:** The author(s) declared no potential conflicts of interest with respect to the research, authorship, and/or publication of this article.

### Competing Interest Statement

The authors have declared no competing interest.

### Author Declarations

The study was conducted in accordance with Nazarbayev University Institutional Research Ethics Committee code of ethics to ensure participant rights and confidentiality

